# Genetically predicted smoking behaviors on Graves’ disease: A two-sample mendelian randomization

**DOI:** 10.1101/2023.10.12.23296814

**Authors:** Mahdi Akbarzadeh, Maryam Pourganji, Arefeh Tabashiri, Sahand Tehrani Fateh, Mahdi Salarabedi, Danial Habibi, Aysan Moeinafshar, Amir Hossein Saeidian, Amir Hossein Ghanooni, Parisa Riahi, Maryam Zarkesh, Hakon Hakonarson, Majid Valizadeh, Fereidoun Azizi, Mehdi Hedayati, Maryam Sadat Daneshpour

**Author notes:** **Corresponding author: Maryam Sadat Daneshpour** (Ph.D.), Associate Professor. Cellular and Molecular Endocrine Research Center, Research Institute for Endocrine Sciences, Shahid Beheshti University of Medical Sciences.; P.O. Box: 19395-4763, 1985717413.

## Abstract

**Background/Aim:** The potential link between smoking and the susceptibility to Graves’ disease (GD) has been scrutinized in observational studies, yielding inconsistent results. We conducted a Mendelian randomization (MR) analysis to ascertain the causal relationship between smoking behaviors and the risk of GD.

**Method:** The data on smoking behaviors, including smoking initiation and lifetime smoking, were obtained from the published GWAS of individuals of European descent who participated in the GSCAN consortium. The genetic variants associated with Graves’ disease were identified using a GWAS of 458,620 participants of European descent from the UK Biobank.

**Results:** Our results show that smoking initiation was associated with GD [OR= 1.50, 95% CI (1.03,2.18), SE□=□0.199, P_beta_□=□0.031; Cochran’s Q=36.62, p=0.999, I^2^=0.0%; MR–Egger_intercept_= 0.003, p= 0.879], and lifetime smoking [OR□=□3.42, 95% CI (1.56, 7.50), SE□=□0.39, P_beta_<0.01; Cochran’s Q=62.68, p=0.99, I^2^=0.0%; Egger_intercept_=0.012, p=0.49]. All other MR methods, as well as sensitivity analysis results, were consistent in terms of betas and significance levels.

**Conclusion:** Our findings lend support to a causal relationship between smoking behaviors and the risk of Graves’ disease. These observations raise important questions about the role of smoking in the progression of GD. So, further investigation is clinically necessary to clarify the links between smoking and GD, which could inform health policy decisions aimed at reducing the risk of GD.

## Introduction

Graves’ disease (GD) is the primary cause of hyperthyroidism in developed countries, with an incidence rate of 20 per 100,000 annually. It is 5-10 times more frequent in women than men, and it peaks at the age of 30-60 (1). GD is an autoimmune disease targeting the thyroid gland by producing antibodies that are agonists of thyroid-stimulating hormone receptor (TSHR), also called anti-TSH receptor antibodies (TRAb). These antibodies cause the overproduction of thyroid hormones that leads to weight loss, increased sweating, heat intolerance, increased appetite, palpitation, tremors, fatigue, and menstrual disorders (2,3). There are also extrathyroidal manifestations, including ophthalmopathy and dermopathy (4). 25-50% of Graves’ patients demonstrate Graves’ ophthalmopathy (5).

The exact etiology of the disease is still unknown. The development of TRAb is caused by breaking down the immune tolerance, which is a multifactorial process in which genetic, epigenetic, and environmental factors are involved (6,7,8). Smoking is one of the main environmental risk factors of GD, especially Graves’ ophthalmopathy (9). Numerous studies have demonstrated a relation between smoking and Graves’ ophthalmopathy, its severity, and poor response to immunosuppressive treatments (10). Studies have suggested that smoking increases reactive oxygen species (ROS) production, which leads to increased adipogenesis and glycosaminoglycans (mostly hyaluronan) synthesis by targeting orbital fibroblasts. This process causes extracellular matrix expansion concomitant with edema (11,12).

Despite all, our data on smoking in association with GD is based on observational studies, which are prone to confounding bias. Therefore, the causal relationship between smoking and GD is still questionable. If the causality is proven, public health interventions aimed at smoking cessation can be adopted to decrease the incidence of the disease. Also, by identifying high-risk individuals, healthcare workers can monitor thyroid function more closely in such cases, which leads to better management of the disease.

Mendelian randomization (MR) is an epidemiological method to minimize the confounding bias in studying the causality between various exposures and outcomes seen in previous observational studies (13). This method relies on the random and independent distribution of genetic variants through miosis (14). In MR, we use genetic variants as instrumental variables; these genetic variants are associated with specific exposures (15). In this study, we use MR design employing European genome-wide association studies (GWAS) information of more than 400,000 participants to investigate the causality between smoking behaviors and GD.

## Methods

### Study setting

In this two-sample Mendelian randomization analysis, summary data from genome-wide association studies (GWAS) have been utilized to identify a causal relationship between smoking behaviors and GD (16). Our analysis aimed to examine the causal effect of smoking behaviors on GD. We used smoking initiation and lifetime smoking as the exposure variables and GD as the outcome variable in this analysis in Figure 1. The summary-level data for each GWAS is detailed in Table 1.

**Table 1:**
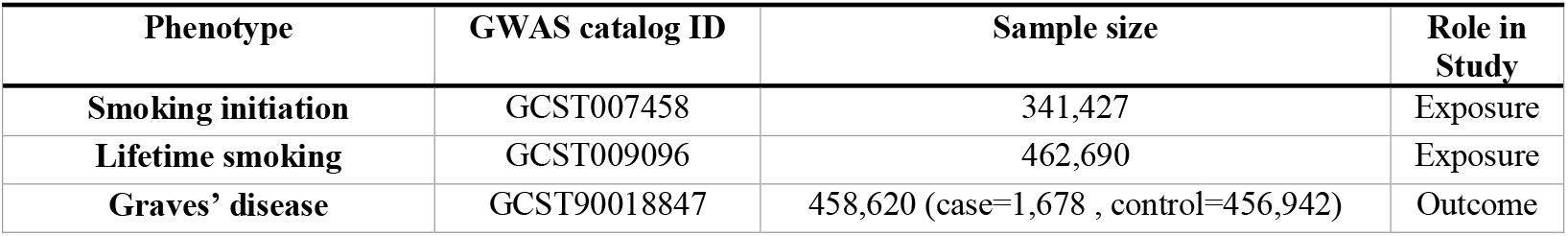
Detailed information of data used for analysis.

| Phenotype | GWAS catalog ID | Sample size | Role in Study |
| --- | --- | --- | --- |
| Smoking initiation | GCST007458 | 341,427 | Exposure |
| Lifetime smoking | GCST009096 | 462,690 | Exposure |
| Graves' disease | GCST90018847 | 458,620 (case=1,678 , control=456,942) | Outcome |

**Figure 1:**
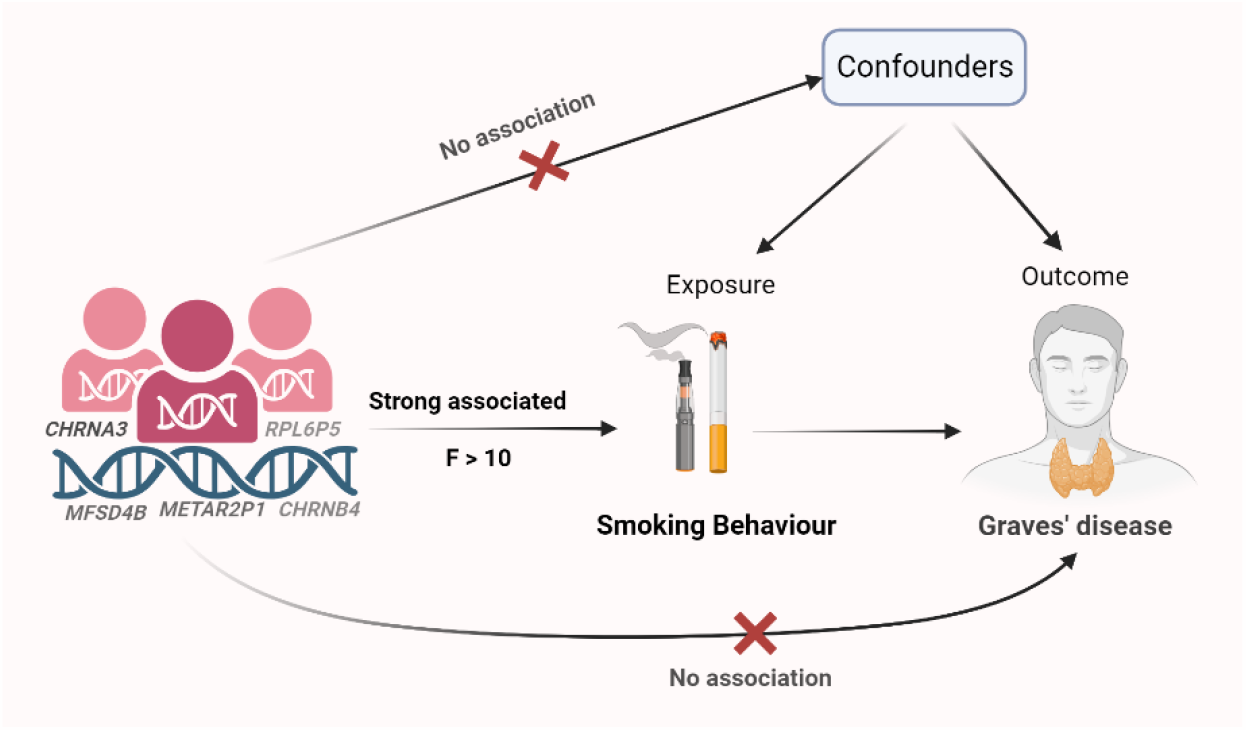
The description of the Mendelian Randomization (MR) assumptions and causal association between smoking behaviors (smoking initiation and lifetime smoking) and risk of Graves’ disease.

The dataset for the GWAS on smoking initiation is available in the GWAS and Sequencing Consortium of Alcohol and Nicotine (GSCAN) (17). The GSCAN investigated smoking behavior initiation using a binary phenotype to determine if an individual had a history of regular smoking. This classification divided individuals into two groups with/without a history of regular smoking (n□=□1,232,091) (17).

In our study, we utilized the data from the GWAS of lifetime smoking behavior conducted by Wootton RE et al. (2020). Lifetime smoking is a comprehensive metric that quantifies the total impact of smoking on an individual’s health throughout their life. It incorporates various aspects of a person’s smoking history, including the duration of smoking(age of initiation), the number of cigarettes they consume per day (cigarettes per day), and the period that has elapsed since they quit smoking (smoking cessation). This measure provides a holistic view of an individual’s exposure to smoking, allowing for a more nuanced understanding of its long-term effects on health. This data was collected from a sample of 462,690 individuals from the UK Biobank (18).

GD is an autoimmune disorder that leads to overproduction of thyroid hormones, known as hyperthyroidism. It’s the most common cause of hyperthyroidism and often results in an enlarged thyroid. In a GWAS conducted on GD, data was obtained from the National Bioscience Database Center (NBDC). The study included 458,620 samples with European ancestry, of which 1,678 individuals identified as having GD and the remaining 456,942 without the disease (19).

### Selection of Instrumental Variables

To validate the assumptions of Mendelian randomization, we performed several quality control steps on the genetic data. Initially, we selected genome-wide significant variants (P□<□5□×□10^−8^) for smoking behaviors from a previous GWAS (20), with an r^2^ value of 0.001, a distance of 10kb, European-ancestry (21), and harmonized them with the summary data on GD to ensure the alleles are correctly aligned. Next, we removed any palindromic SNPs and any SNPs with low minor allele frequency (MAF) (less than 0.01), as they may introduce statistical bias or low confidence in the original GWAS (22). Then, we excluded any SNPs that were not present in the outcome dataset and used proxy SNPs with high linkage disequilibrium (LD) (r^2^ greater than 0.8) from the NIH LD link website considering European (EUR) ancestry (23). Finally, we calculated the F-statistic for each instrumental variable (IV) to measure its strength and excluded any IVs with F-statistic lower than 10, as they may be weak instruments and bias the estimates (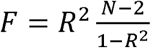, where 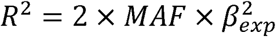 where *R*^2^ is the total variance of the extracted SNPs, and N is the sample size) (24). These steps ensured that the first two assumptions of Mendelian randomization, namely the relevance and independence of the IVs, were met. However, the third assumption, the exclusion restriction assumption, was not directly tested in our analysis, as it is often hard to verify empirically. Therefore, we performed sensitivity analyses to assess the robustness of our results to potential violations of this assumption, such as pleiotropy or mediation effects. We used different methods such as MR-Egger regression (intercept), Phenoscanner tools, and MR-PRESSO to estimate the causal effect of smoking on GD under different scenarios.

### Two-sample MR analysis

The inverse variance-weighted (IVW) approach was employed as the primary method for calculating the aggregate effect of all single nucleotide polymorphisms (SNPs). A P-value below 0.05 was considered statistically significant after the Mendelian randomization, which was conducted using *TwoSampleMR, MendelianRandomization*, and *mr*.*raps* packages in R (version 4.0.3) (25). Two-sample MR is susceptible to heterogeneity due to variations in analysis platforms, populations, and experimental conditions. We employed Cochran’s Q statistic and the I^2^ index for MR-IVW analyses to detect heterogeneity and Rucker’s Q statistic for MR-Egger (26). In addition, The Egger bias intercept test was employed to detect the presence of horizontal pleiotropy quantitatively (27). Diagnostic leave-one-out, single-nucleotide variant, and funnel-plot analyses were conducted to identify outliers and bias(28,29). We also consulted the human gene phenotypic association database (PhenoScanner) to identify and remove pleiotropic SNPs directly associated with outcome or confounder variables (30,31). The MR-PRESSO test was conducted to detect potential outliers and correct for horizontal pleiotropy through outlier removal, which was conducted using the *MRPRESSO* package in R. Cook’s distance was utilized to identify and remove SNPs that had a strong and false influence on the model’s fitted values (32). DFFITS, Standardized residuals, Studentized residuals, and Studentized Residuals vs. Leverage Plot were employed to determine if any individual SNPs were detected as outliers or influential points in the MR analysis (33). To assess the validity and robustness of our results, we conducted several sensitivity analyses, including the MR-Egger, weighted median, simple mode, and weighted mode methods.

To ensure that the primary assumptions of the MR analysis were not violated, we conducted a thorough sensitivity analysis including Maximum Likelihood Estimation (MLE) method, Robust Adjusted Profile Score (RAPS), MR-Lasso, constrained maximum likelihood (MR-cML), mode-based, and debiased inverse variance weighted methods. A 2-sided P-value□<□0.05 was used as the criterion for statistical significance in the analysis of the effect of smoking on GD outcomes. The relative associations of the GD outcomes with lifetime smoking and smoking initiation were assessed by the odds ratio (OR) and its 95% confidence interval (CI).

## Results

Detailed findings related to lifetime smoking components, that is, the age of initiation, the number of cigarettes smoked per day, and any attempts at smoking cessation, are provided in the supplementary file accompanying this paper. We strongly recommend that readers should refer to this supplementary material for a comprehensive understanding of all the exposures studied. The results for smoking initiation and lifetime smoking are detailed in this section.

### SNPs selection

The GWAS for lifetime smoking and smoking initiation identified 7,846 and 10,413 SNPs that reached the genome-wide significance level (P < 5×10^−8^), respectively. After clumping those SNPs, 93 and 126 independent SNPs remained for MR analysis. Those SNPs were further used as outcomes in the harmonization step for GD. We performed the harmonization step (action=2) after removing SNPs that met the following criteria: palindromic, sourced from the PhenoScanner tool or classified as outliers or influential points. The harmonization step resulted in 84 SNPs for smoking initiation and 115 SNPs for lifetime smoking (Supplementary Table S1 and S2).

### The causal relationship between smoking initiation and GD

The application of the IVW method revealed the potential existence of a positive association between smoking initiation and GD. It shows that an increase in smoking initiation is associated with an increased risk of GD (OR= 1.50, 95% CI (1.03,2.18), SE□=□0.199, P_beta_□=□0.031). The analysis found no evidence of pleiotropy (MR–Egger intercept = 0.003, P = 0.879) or heterogeneity (Cochran’s Q=36.62, P=0.999, I^2^=0.0%;). This analysis was based on 65 SNPs that passed the quality control criteria and were used as instrumental variables for smoking initiation.

### The causal relationship between lifetime Smoking and GD

We next considered the effect of lifetime smoking on GD. Findings showed a significant association between lifetime smoking and an increased risk of GD (OR□=□3.42, 95% CI (1.56, 7.50), SE□=□0.39, P_beta_□<□0.01). Evaluating the heterogeneity and pleiotropy test showed Cochran’s Q= 62.68, p=0.99 I^2^=0.0%; Egger intercept=0.012, P=0.49; hence, no evidence of heterogeneity or pleiotropy was found. This analysis was based on 95 SNPs that passed the quality control criteria and were used as instrumental variables for lifetime smoking.

### Sensitivity analyses

The MR pleiotropy residual sum and outlier test (MR-PRESSO), along with the MR pleiotropy residual sum and outlier (Radial MR) and Cook’s distance, were employed to evaluate the potential existence of outlying SNPs and to reevaluate the effect estimates (Supplementary Figure S1).

Scatter plots (Supplementary Figure S2) and leave-one-out plots (Supplementary Figure S4) assessed the impact of outlying values. In addition, funnel and forest plots were used to illustrate the causal relationship between smoking and GD (Supplementary Figure S6).

**Figure 1.**
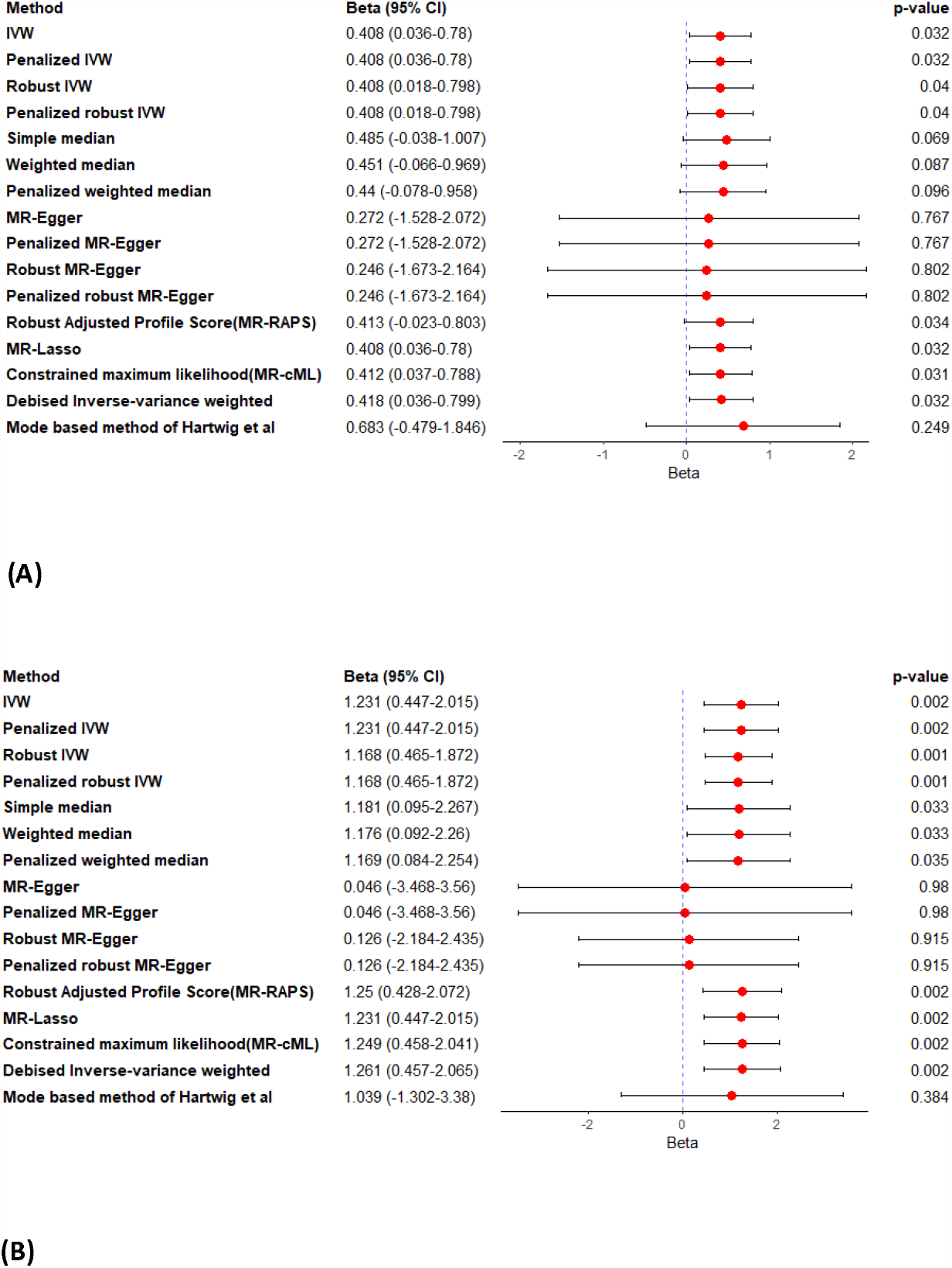
Figure A shows the different methods used to estimate the effect of starting to smoke on Graves’ disease in a Mendelian randomization analysis. Figure B does the same for lifetime smoking. The IVW method assumes that the genetic variants are not related, while the Egger method allows for some relationship between the variants. The confidence intervals are shown at a 95% level.

**Figure 2.**
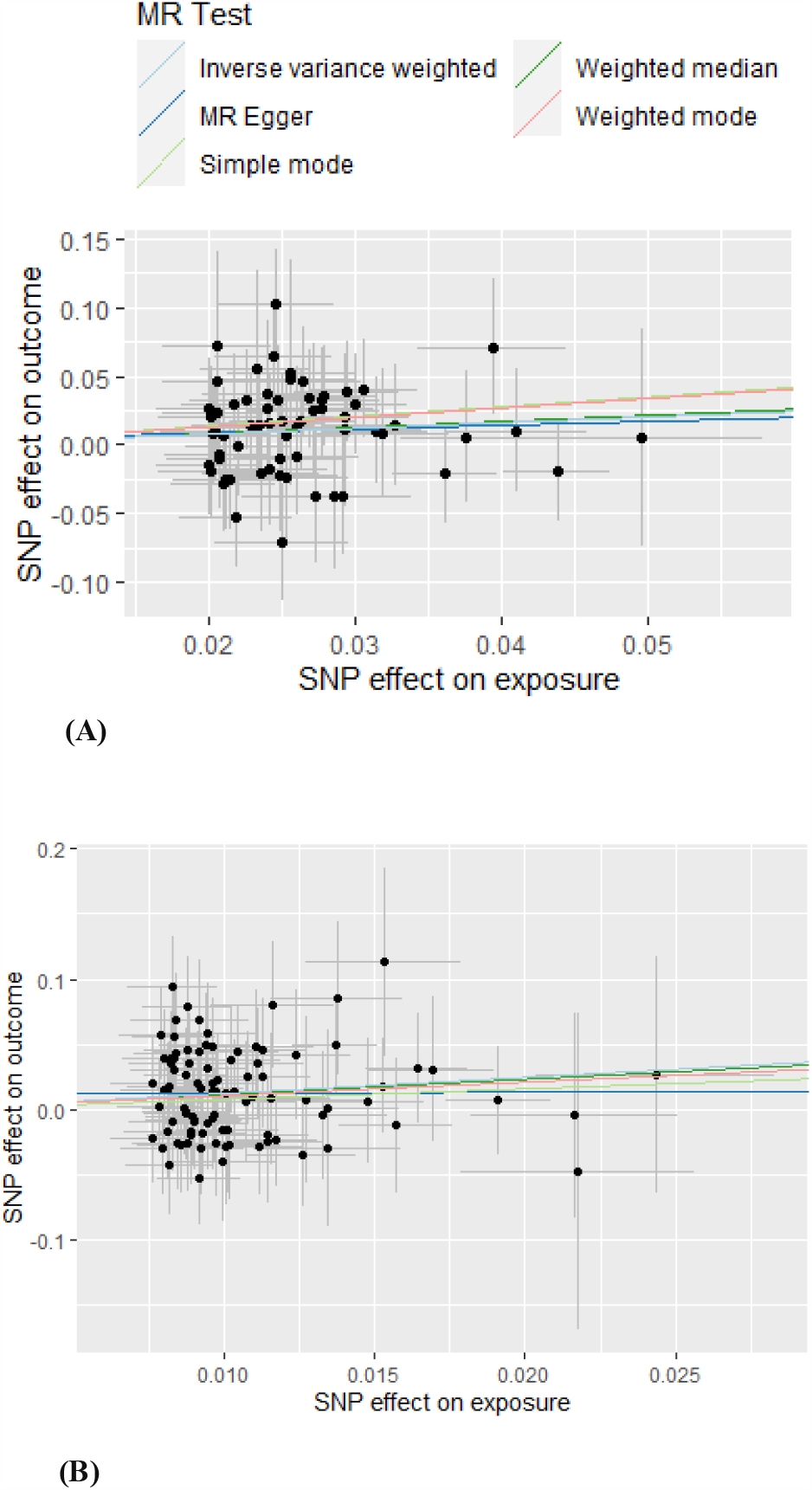
Figures A and B present scatter plots, illustrating the results obtained using the main methods, including inverse variance weighted, weighted median, MR Egger, weighted mode, and simple mode. Figure A represents smoking initiation, and Figure B represents lifetime smoking.

## Discussion

To the best of our knowledge, the present study is the first study to examine the causal relationship between cigarette smoking and GD by using a two-sample MR method. Here, we used GWAS data to investigate GD in individuals who are genetically predisposed to smoking compared to controls. We performed various MR analyses using loci associated with different stages of cigarette use, including smoking initiation, heaviness (cigarettes per day), and age of initiation. Our findings suggested that there is a causal association between smoking initiation and lifetime smoking on GD.

Due to the harmful nature of smoking, it is not ethically possible to conduct randomized clinical trials in this regard. Therefore, our previous knowledge about the association between smoking and GD was limited to observational studies. In 1993, Prummel et al. conducted a case-control study to assess the smoking status in five groups: GD with and without ophthalmopathy, toxic and nontoxic goiter, and autoimmune hypothyroidism. The results of their study showed that smoking significantly increased Graves’ ophthalmopathy with an odds ratio of 7.7 (95% CI, 4.3-13.7). However, this strong association was not detected in GD without ophthalmopathy (OR, 1.9; 95% CI, 1.1-3.2) (34). In 1998, Yoshiuchi et al. investigated the role of smoking in GD in men and women independently. They adjusted data for stressful life events, drinking habits, coping skills, social support, and daily hassles. Finally, their results demonstrated a significant association between smoking and GD in women (95% CI, 2.2-27; p-value<0.001). However, this association was not seen in men (35). In 2002, Vestergaard performed a meta-analysis of eight available studies. The risk of GD was 3.3 (95% CI, 2.09-5.22) times higher in current smokers than controls. However, there was no excess risk of GD in ex-smokers (OR, 1.41; 95%CI, 0.77-2.58). It is worth noting that smoking had a significantly stronger association with Graves’ ophthalmopathy (OR, 4.4; 95%CI, 2.88-6.73) compared to GD (OR, 1.95; 95%CI, 1.42-2.55) (36). In 2006, Thornton et al. conducted another systematic review and meta-analysis of 15 studies on smoking and thyroid ophthalmopathy. This study demonstrated a positive association between smoking and Graves’ ophthalmopathy (OR, 1,22-20.2) in case-control studies where the control subjects had no thyroid disease. This positive association was also seen in studies that compared Graves’ ophthalmopathy with Graves’ cases without ophthalmopathy (OR, 1.94-10.1). They also reported poorer outcomes of thyroid eye disease in smokers (37). In contrast to the mentioned studies, a large population-based cross-sectional study suggested a negative association between smoking and thyroid autoantibodies; however, it was associated with mildly decreased TSH levels (38).

According to the above studies, it can be concluded that there is not much difference between the studies regarding the relationship between smoking and Graves’ ophthalmopathy. In contrast, the findings of the studies regarding the relationship between smoking and GD in overall are somewhat contradictory. On the other hand, the existing genome-wide association studies had very limited populations in regard to Graves’ ophthalmopathy. Therefore, we decided to investigate the relationship between smoking and GD itself and not the ophthalmopathy caused by it.

In addition to the observational studies mentioned above, a number of studies have attempted to elucidate the possible molecular mechanisms explaining this association. However, most of the molecular studies focused on the effect of smoking on fibroblasts and Graves’ ophthalmopathy. Kau et al. investigated the effect of cigarette smoke extract on some intracellular mechanisms of fibroblasts. They showed that smoking decreases cell viability by inducing oxidative stress in a dose-dependent manner. It also induces the levels of fibrosis-related proteins and gene expression in fibroblasts (39). Görtz et al. addressed this issue from another aspect. They demonstrated that smoking increases hypoxia-inducible factor-1 (HIF-1), leading to the increased secretion of vascular endothelial growth factor (VEGF), increased adipogenesis, and enhanced adiponectin release, all contributing to tissue enlargement (40).

In this MR study, exposure variables were smoking-related genetic variants, including lifetime smoking, cigarettes per day, age of initiation, smoking initiation, and smoking cessation. Lifetime smoking is an index reflecting smoking duration, cessation, and smoking heaviness (18). Since it reflects various aspects of smoking, it is potentially more prone to horizontal pleiotropy (41). Despite conducting sensitivity analysis, this issue justifies our use of other exposure variables in addition to lifetime smoking. Our study suggests a causal relationship between all exposure variables except smoking cessation and GD. Although all of the mentioned causalities are significant, not all of them are strong.

Among the five smoking mentioned above status traits, two were dichotomous: smoking initiation and cessation. Based on our analysis, smoking initiation was significantly associated with GD (beta□=□0.408, 95% CI (0.02,0.78); SE□=□0.199, P_beta_□=□0.031), meaning that lifetime smoking exposure has a significant effect on disease incidence. This finding of our study is consistent with the previous studies conducted on the association between lifetime smoking exposure and the occurrence of GD (beta□=□1.23, 95% CI (0.45, 2.01); SE□=□0.39, P_beta_□=□0.002). However, our analysis showed no statistically significant association between smoking cessation and GD (beta□=□-0.14, 95% CI (1.06,1.34); SE□=□0.60, P_beta_□=□0.81). Although other previous studies have mentioned no excess risk of GD in ex-smokers, this finding shows that smoking cessation does not capture the association by itself.

Among the strengths of this study, the following can be mentioned: I) This is the first study to investigate the causal relationship between smoking and GD using the MR method. In this method, the effect of confounders is minimized due to using genetic variants as IVs and random distribution of genetic variants through miosis. II) We used a reliable GWAS database and a large sample size for our analysis. III) We employed multiple methods to detect pleiotropy to reduce the bias; IV) Finally, we analyzed all of the smoking traits (lifetime smoking, cigarettes per day, age of initiation, smoking initiation, and smoking cessation) as exposure variables to investigate any association.

The main limitations of our study include: I) We selected IVs from a European descent GWAS, which limits the generalizability of the study to other populations. II) Similar to all of the MR studies, it is possible that horizontal pleiotropy still exists despite different tests to detect it. III) Small beta suggests that the causalities are not strong, meaning that various known or unknown factors are involved. Therefore, the interpretation of the results should be done with caution.

## Conclusions

Findings from our MR analysis support a causal link between smoking behaviors and the risk of various GD outcomes. However, these conclusions should be interpreted with caution due to the inherent limitations of MR studies and the intricate nature of the relationship between smoking behaviors and the risk of various GD. Further research employing more precise methodologies and measurement tools in populations with diverse characteristics is necessary to corroborate the evidence derived from this study. Additionally, conducting clinical studies could shed light on the underlying biological mechanisms that justify the observed positive causal relationship between smoking behaviors and the risk of various GD.

## Declarations

## Acknowledgments

We want to acknowledge the participants and investigators who made summary data available.

## Consent for publication

Not applicable.

## Data Availability Statement

The original data used are publicly available at https://www.ebi.ac.uk/gwas/. The original contributions presented in the study are included in the article/Supplementary Material. The process of MR analyses and the results are publicly available through the following HTML link: https://akbarzadehms.github.io/SmokinGravesMR/

## Ethical approval and consent to participate

The ethical committee approved this study at the Research Institute for Endocrine Sciences, Shahid Beheshti University of Medical Sciences (Research Approval Code: 28778 & Research Ethical Code: IR.SBMU.ENDOCRINE.REC.1400.084). In this study, all participants provided written informed consent for participating in the study. This study has been performed following the Declaration of Helsinki.

## Author Contributions

MA and STF conceptualized the study; MA and MP analyzed the data. MA, MP, and AT drafted the initial manuscript, generated Tables and Figures, and designed the HTML file. MS, DH, AM, AHS, and AHG confirmed the results. PR, MZ, HH, MV, FA, MH, and revised the manuscript. MSD supervised, edited, and finalized the manuscript. All authors reviewed and approved the final manuscript.

## References

1. Antonelli A Fsrfegpsri et al. Graves’ disease: Epidemiology, genetic and environmental risk factors and viruses. Best Practice & Research Clinical Endocrinology & Metabolism. 2020;34(1).

2. Ehlers M SMAS. Graves, disease in clinical perspective. Frontiers in Bioscience-Landmark. 2019;24(1).

3. Davies TF Aslrnybgbm et al. Graves’ disease. Nature reviews Disease primers. 2020;6(1).

4. Smith TJ HL. Graves’ disease.. New England Journal of Medicine. 2016;375(16).

5. Bahn RS. Graves’ ophthalmopathy. N Engl J Med. 362(8).

6. Pokhrel B BK. Graves disease. 2017.

7. Wémeau J-L Kmsjlbcvcfl editors. Graves’ disease: introduction, epidemiology, endogenous and environmental pathogenic factors. Annales d’endocrinologie. Elsevier. 2018;

8. Razmara E Smasbayhcj et al. Graves’ disease: introducing new genetic and epigenetic contributors. Journal of molecular endocrinology. 2022;66(2).

9. Bufalo NE Srcaarmjme et al. Genetic polymorphisms associated with cigarette smoking and the risk of Graves’ disease. Clin Endocrinol (Oxf). 2008;68(6).

10. Thornton J Kshrer. Cigarette smoking and thyroid eye disease: a systematic review. Eye. 2007;21(9).

11. Fallahi P Fsegrfpspa et al. Cytokines as targets of novel therapies for Graves’ ophthalmopathy. Frontiers in endocrinology. 2021;

12. Aranyosi JK Geeakmfmuz et al. Different effects of cigarette smoke, heated tobacco product and e-cigarette vapour on orbital fibroblasts in graves’ orbitopathy; A study by real time cell electronic sensing. Molecules. 2022;27(9).

13. Richmond RC SG. Mendelian randomization: concepts and scope. Cold Spring Harbor perspectives in medicine. 2022;12(1).

14. Emdin CA Kaks. Mendelian randomization. JAMA. 2017;318(19).

15. Sanderson E Gmhmkhmjmm et al. Mendelian randomization. Nature Reviews Methods Primers. 2022;2(1).

16. Haycock PC Bmbklrhsbs et al. Design and quality control of large-scale two-sample Mendelian randomisation studies. medRxiv. 2021;

17. Liu M, Jiang Y, Wedow R, Li Y, Brazel DM, Chen F, et al. Association studies of up to 1.2 million individuals yield new insights into the genetic etiology of tobacco and alcohol use. Nat Genet [Internet]. 2019;51(2):237–44. Available from: 10.1038/s41588-018-0307-5

18. Wootton RE Rrsblrshtg et al. Evidence for causal effects of lifetime smoking on risk for depression and schizophrenia: a Mendelian randomisation study. Psychol Med. 2020;50(14).

19. Sakaue S et al. A cross-population atlas of genetic associations for 220 human phenotypes. Nat Genet. 2021;35(10).

20. Tylee DS Lywfpglddaf et al. An atlas of genetic correlations and genetically informed associations linking psychiatric and immune-related phenotypes. JAMA Psychiatry. 2022;79(7):667–76.

21. Marees AT de KHSSVFCEMC et al. A tutorial on conducting genome-wide association studies: Quality control and statistical analysis. Int J Methods Psychiatr Res. 2018;27(2).

22. Cui Z TY. Using genetic variants to evaluate the causal effect of serum vitamin D concentration on COVID-19 susceptibility, severity and hospitalization traits: a Mendelian randomization study. J Transl Med. 2021;19(1).

23. https://ldlink.nih.gov/?tab=ldproxy.

24. Bowden J Dgmfmczqldsn et al. Improving the accuracy of two-sample summary-data Mendelian randomization: moving beyond the NOME assumption. Int J Epidemiol. 2019;48(3).

25. Nikolakopoulou A Mdsg. How to interpret meta-analysis models: fixed effect and random effects meta-analyses. Evid Based Ment Health. 2014;17(2).

26. Hemani G Zjebwkhvbd et al. The MR-Base platform supports systematic causal inference across the human phenome. Elife. 2018;

27. Burgess S TS. Interpreting findings from Mendelian randomization using the MR-Egger method. Eur J Epidemiol. 2017;32(5).

28. Burgess S Sgdndfgdgm et al. Guidelines for performing Mendelian randomization investigations. Wellcome Open Res. 2019;

29. Bowden J DSGBS. Mendelian randomization with invalid instruments: effect estimation and bias detection through Egger regression. Int J Epidemiol. 2015;44(2).

30. Kamat MA Bjyrspbsdj et al. PhenoScanner V2: an expanded tool for searching human genotype–phenotype associations. Bioinformatics. 2019;35(22).

31. Staley JR Bjkmesspsb et al. PhenoScanner: a database of human genotype–phenotype associations. Bioinformatics. 2016;32(20).

32. Burgess S TS. Interpreting findings from Mendelian randomization using the MR-Egger method. Eur J Epidemiol. 2017;32(5):377–89.

33. Burgess S Dndfgdgmhf et al. Guidelines for performing Mendelian randomization investigations: update for summer 2023. 2023;

34. Prummel MF WW. Smoking and Risk of Graves’ Disease. JAMA. 1993;269(4).

35. Yoshiuchi K Khnsyhikky et al. Stressful life events and smoking were associated with Graves’ disease in women, but not in men. Psychosom Med. 1998;60(2).

36. Vestergaard P. Smoking and thyroid disorders–a meta-analysis. Eur J Endocrinol. 2002;146(2).

37. Thornton J Kshrer. Cigarette smoking and thyroid eye disease: a systematic review. Eye. 2007;21(9).

38. Belin RM ABPNLP. Smoke exposure is associated with a lower prevalence of serum thyroid autoantibodies and thyrotropin concentration elevation and a higher prevalence of mild thyrotropin concentration suppression in the third National Health and Nutrition Examination Survey (NHANES III). J Clin Endocrinol Metab. 2004;89(12).

39. Kau H-C Wsbtcclclwyh. Cigarette smoke extract-induced oxidative stress and fibrosis-related genes expression in orbital fibroblasts from patients with Graves’ ophthalmopathy. Oxid Med Cell Longev. 2016;

40. Görtz G-E Hmabrbfjea et al. Hypoxia-dependent HIF-1 activation impacts on tissue remodeling in Graves’ ophthalmopathy—implications for smoking. The Journal of Clinical Endocrinology & Metabolism. 2016;101(12).

41. Park HA Nsmkbmwqdj et al. Mendelian randomisation study of smoking exposure in relation to breast cancer risk. British journal of cancer. 2021;125(8).

