## Supplementary material for "Genetically predicted smoking behaviors on Graves’ disease: A two-sample mendelian randomization": supplemantry figures

Supplementary materials

**A cause-effect relationship between Graves’ disease and the smoking behaviours: A two-sample Mendelian randomization study**

**Authors:**

1. Mahdi Akbarzadeh; Ph.D., Cellular and Molecular Endocrine Research Center, Research Institute for Endocrine Sciences, Shahid Beheshti University of Medical Sciences, Tehran, Iran..
2. Maryam Pourganji, MS.c., Cellular and Molecular Endocrine Research Center, Research Institute for Endocrine Sciences, Shahid Beheshti University of Medical Sciences, Tehran, Iran,
3. Arefeh Tabashiri, M.D., Student Research Committee, School of Medicine, Shahid Beheshti University of Medical Sciences, Tehran, Iran..
4. Sahand Tehrani Fateh; MD, School of Medicine, Tehran University of Medical Sciences, Tehran, Iran.
5. Mahdi Salarabedi; M.D., Student Research Committee, School of Medicine, Shahid Beheshti University of Medical Sciences, Tehran, Iran.
6. Danial Habibi; Ph.D., Department of Biostatistics and Epidemiology, School of Health, and Student Research Committee, School of Health, Isfahan University of Medical Sciences, Isfahan, Iran.
7. Aysan Moeinafshar, MD, School of Medicine, Tehran University of Medical Sciences, Tehran, Iran.
8. Amir Hossein Saeidian, Ph.D., Center for Applied Genomics, The Children’s Hospital of Philadelphia, Abramson Research Building, Suite 1016I, 3615 Civic Center Boulevard, Philadelphia, PA, 19104-4318, USA,
9. Amir Hossein Ghanooni; MD, Department of Endocrinology, School of Medicine, Iran University of Medical Sciences, Tehran, Iran.
10. Parisa Riahi; MSc, Cellular and Molecular Endocrine Research Center, Research Institute for Endocrine Sciences, Shahid Beheshti University of Medical Sciences, Tehran, Iran.
11. Maryam Zarkesh: Cellular and Molecular Endocrine Research Center, Research Institute for Endocrine Sciences, Shahid Beheshti University of Medical Sciences, Tehran, Iran.
12. Hakon Hakonarson, MD, PhD, Center for Applied Genomics, The Children’s Hospital of Philadelphia, Abramson Research Building, Suite 1016I, 3615 Civic Center Boulevard, Philadelphia, PA, 19104-4318, USA, Department of Pediatrics, University of Pennsylvania Perelman School of Medicine, Philadelphia, PA, USA, Division of Human Genetics, The Children’s Hospital of Philadelphia, Philadelphia, PA, USA,
13. Majid Valizadeh, MD, Obesity Research Center, Research Institute for Endocrine Sciences, Shahid Beheshti University of Medical Sciences, Tehran, Iran
14. Fereidoun Azizi; MD, Endocrine Research Center, Research Institute for Endocrine Sciences, Shahid Beheshti University of Medical Sciences, Tehran, Iran..
15. Mehdi Hedayati; Ph.D., Cellular and Molecular Endocrine Research Center, Research Institute for Endocrine Sciences, Shahid Beheshti University of Medical Sciences, Tehran, Iran..
16. Maryam Sadat Daneshpour*; Ph.D., Cellular and Molecular Endocrine Research Center, Research Institute for Endocrine Sciences, Shahid Beheshti University of Medical Sciences, Tehran, Iran..

**Corresponding author:**

**Maryam Sadat Daneshpour** (Ph.D.), Associate Professor. Cellular and Molecular Endocrine Research Center, Research Institute for Endocrine Sciences, Shahid Beheshti University of Medical Sciences.; P.O. Box: 19395- 4763, 1985717413, Tel: +98 (21) 22432500, Fax: +98 (21) 22402463.

### **Figure S1. The investigating of outliers and potential SNPs.**

MR-PRESSO and RadialMR did not identify any potential hotspots, but Cook's distance could identify SNPs that affected the results. The below figure shows the results.

**Smoking initiation**

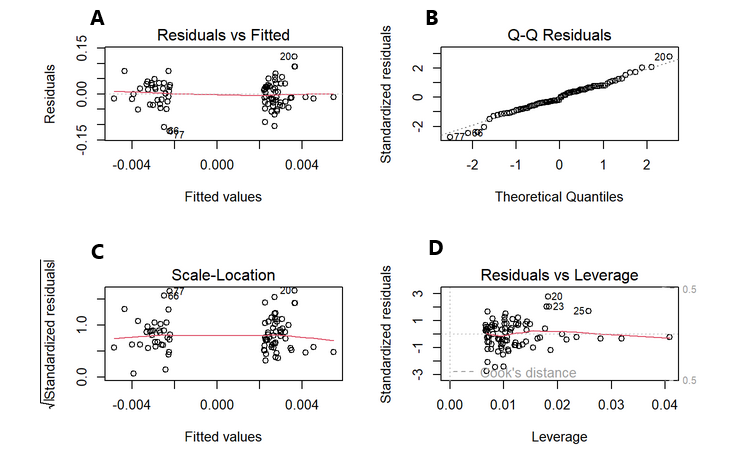

Figure B shows the q-q plot to detect the outlier, and we identified three SNPs (rs9423279, rs12027999, rs7631735).Figure A depicts the relationship between the residuals and the fitted values and Figure C, displays standardized residuals they identify the same SNPs as Figure A (rs9423279, rs12027999, rs7631735).Figure D demonstrates the Cook's distance, and we here realized two SNPs )rs12042107 and rs72789632(, which potential influential point could affect the results.

**Lifetime Smoking**

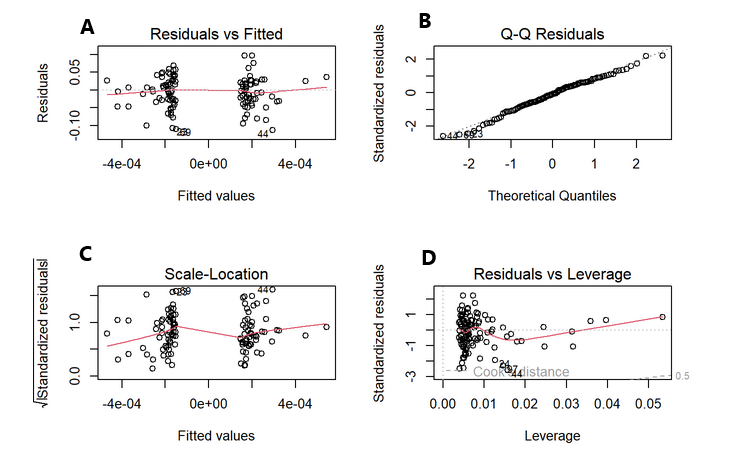

Figure A illustrates the correlation between residuals and fitted values. Figure B presents a Q-Q plot utilized for outlier detection. Figure C exhibits standardized residuals, identifying the same SNPs (rs57611503, rs13009008, rs8042849). Figure D displays Cook’s distance, where we identified two SNPs (rs7077678 and rs329120) that could potentially be influential points affecting the results.

**Cigarettes per day**

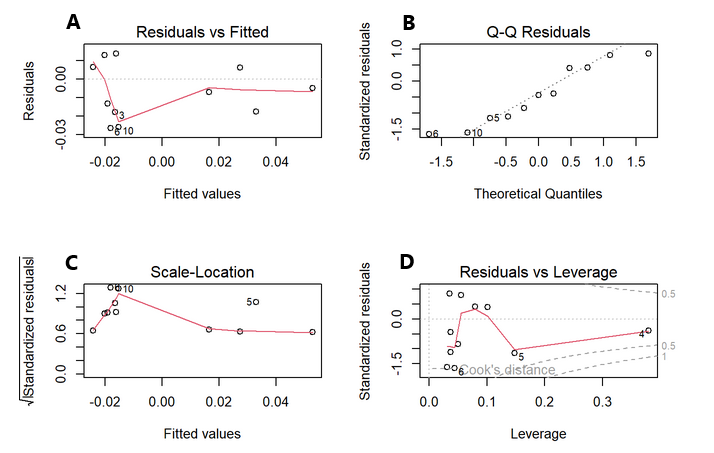

Figure A illustrates the correlation between residuals and fitted values. Figure B presents a Q-Q plot utilized for outlier detection. Figure C exhibits standardized residuals and Figure D displays Cook’s distance, where we identified one SNPs (rs56113850) that could potentially be influential points affecting the results.

**Age of initiation smoking**

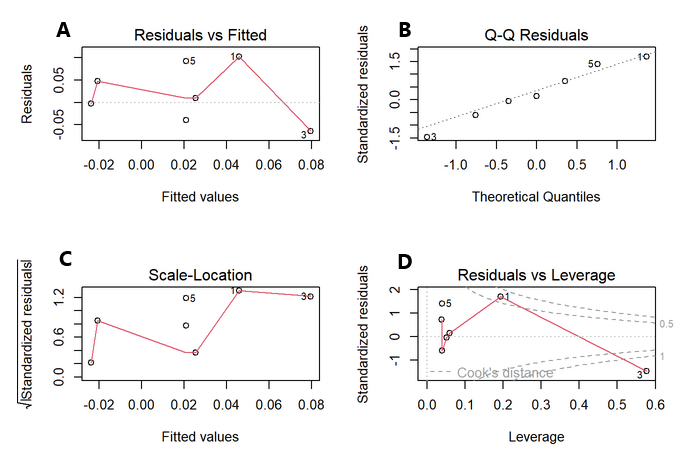
Figure A illustrates the correlation between residuals and fitted values. Figure B presents a Q-Q plot utilized for outlier detection. Figure C exhibits standardized residuals and Figure D displays Cook’s distance, where we identified one SNP (rs11780471) that could potentially be influential points affecting the results.

### **FigureS2. Comparison of the causal estimates from the various Mendelian randomization methods and scatter plot of the potential effects of Smoking behaviors SNPs on Graves’ diseases**

Figures below presents a comparative analysis of the causal estimates derived from multiple Mendelian randomization methodologies. The plot demonstrates a positive causal relationship between Smoking initiation, lifetime smoking, cigarettas per day, age of initiation smoking and smoking cessation on the incidence of Graves’ disease.

**Smoking Initiation**

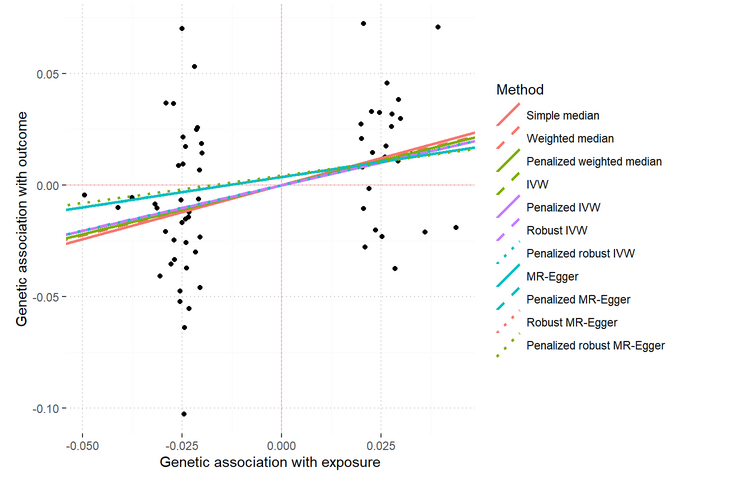

**Lifetime smoking**

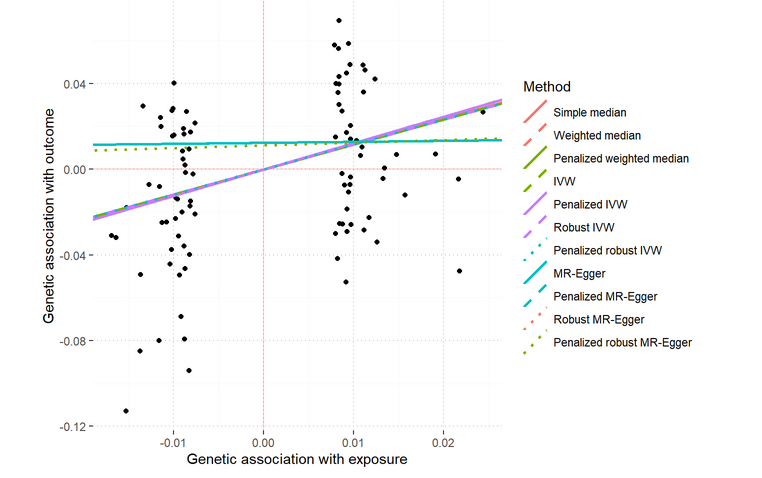

**Cigarettes per day**

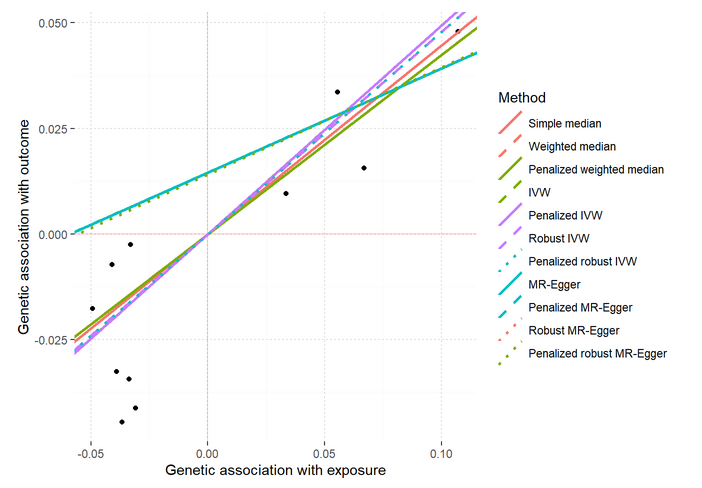

**Age of initiation smoking**

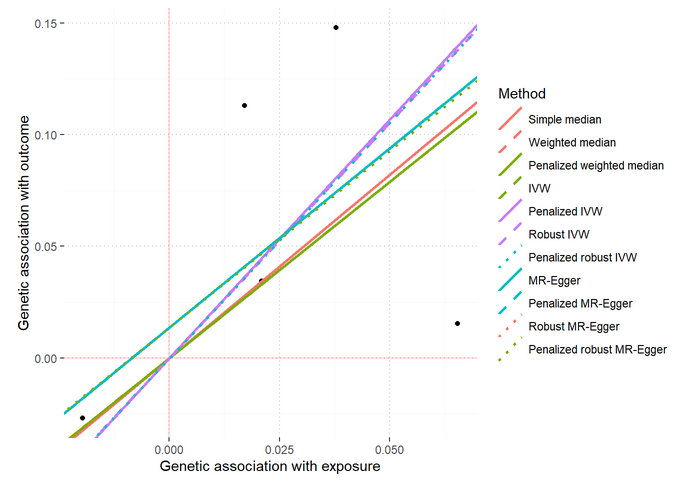

**Smoking cessation**

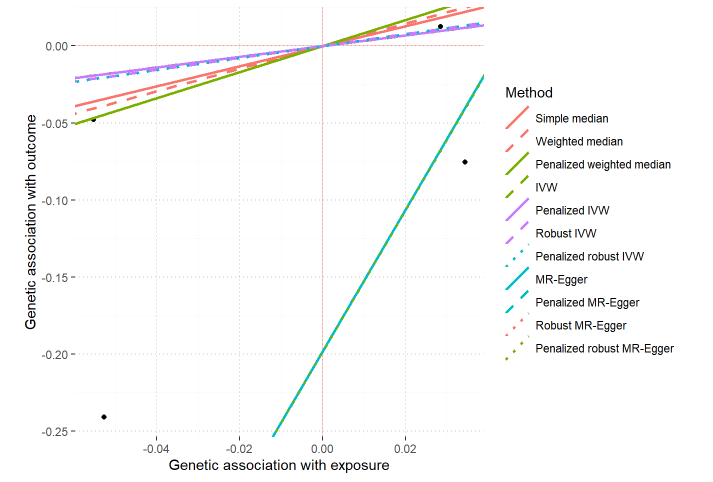

### **Figure S3. scatter plots for the potential impact of SNPs**

Figures presents scatter plots illustrating the potential impact of SNPs associated with smoking initiation and lifetime smoking, cigarettas per day, age of initiation smoking and smoking cessation on the incidence of Graves’ disease. The x-axis represents the genetic association of both exposure variables, while the y-axis represents the genetic association of the outcome variable. The dashed line corresponds to the results obtained using the inverse variance weighted method, and exhibits a positive slope, indicating a positive relationship between the exposure and outcome variables.

**Smoking Initiation**

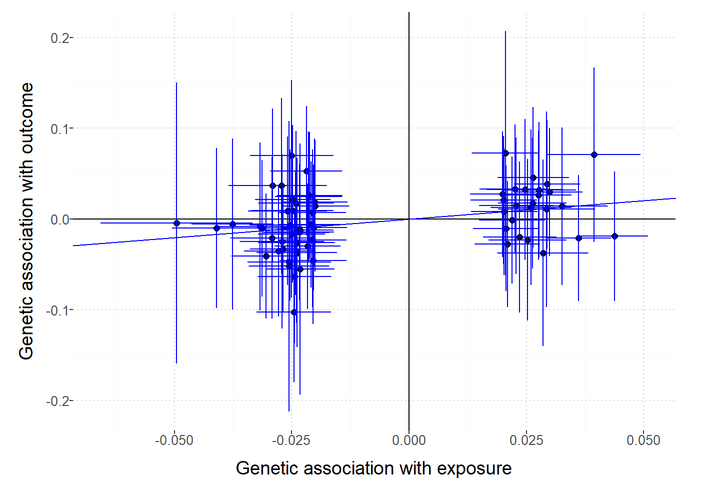

**Lifetime smoking**

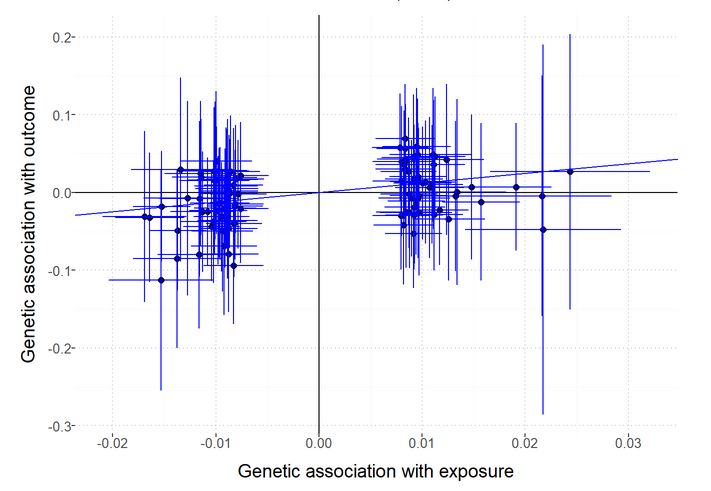

**Cigarettes per day**

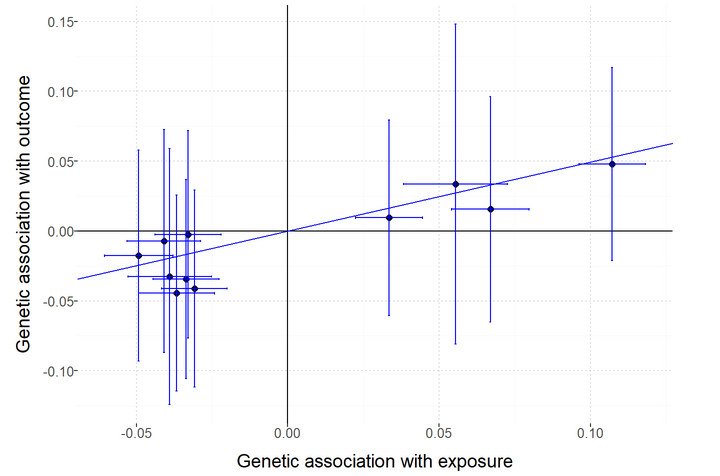

**Age of initiation smoking**

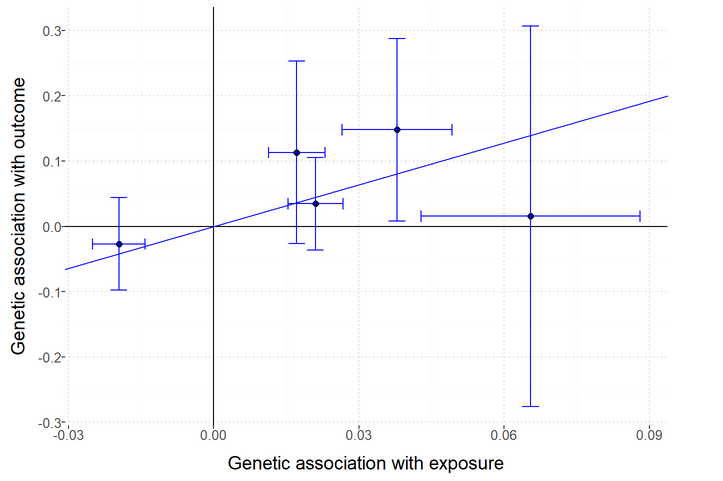

**Smoking cessation**

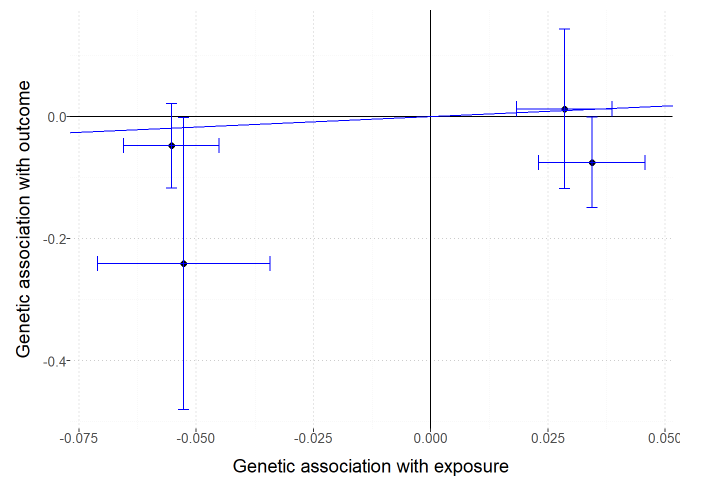

### **Figure S4. Leave-one-out plot to assess if a single variant is driving the association between Smoking behaviors and Graves’ diseases**

Figure S4 presents a leave-one-out analysis to evaluate the influence of individual variants on the observed association between smoking initiation, lifetime smoking, cigarettas per day, age of initiation smoking and smoking cessation on the incidence of Graves’ disease. Effect estimates are reported per SD increase in the exposure and error bars represent 95% confidence intervals.

**Smoking Initiation**

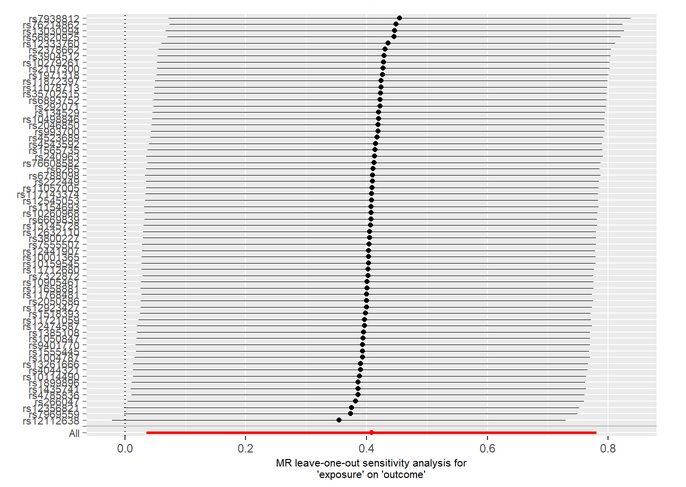

Leave one out plot demonstrating no outliers in MR analysis of smoking initiation (*n=* 341,427) on GD (*n=* 458,620).

**Lifetime smoking**

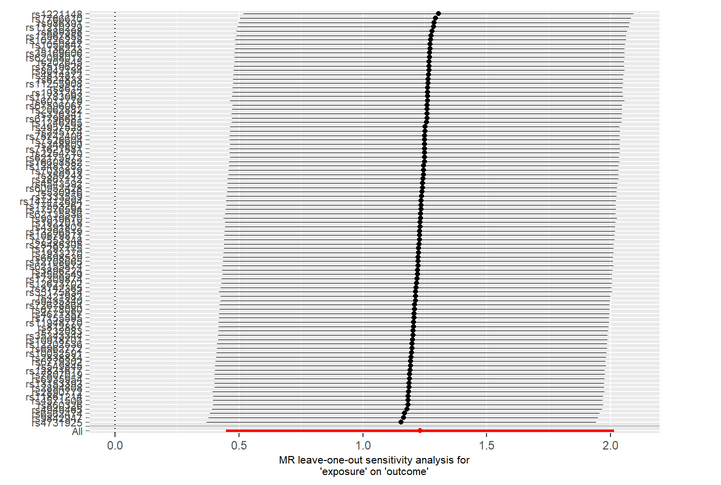

Leave one out plot demonstrating no outliers in MR analysis of and lifetime smoking(n=462,690 ) on GD (n= 458,620).

**Cigarettes per day**

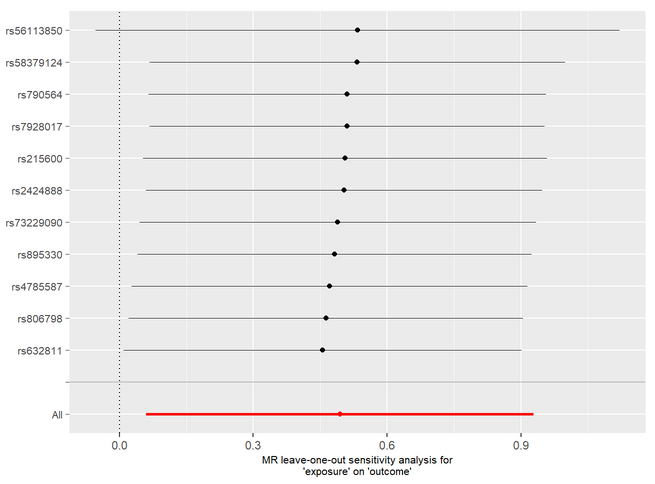

Leave one out plot demonstrating no outliers in MR analysis Cigarettes per day (n= 377,334) on GD (n= 458,620).

**Age of initiation smoking**

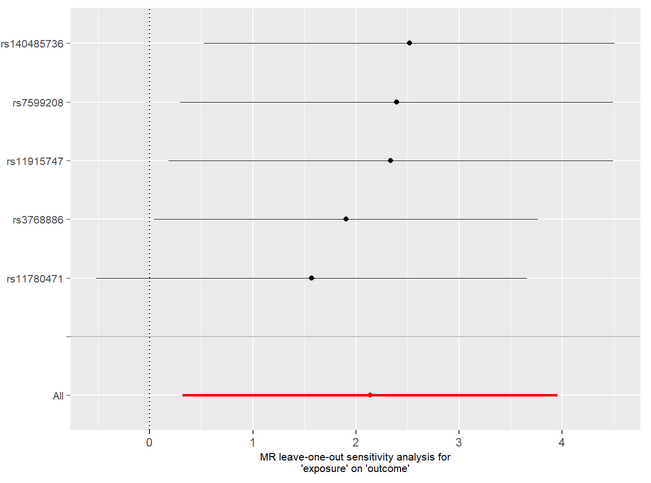

Leave one out plot demonstrating no outliers in MR analysis of Age of initiation smoking (n=341,427) on GD (n= 458,620).

**Smoking cessation**

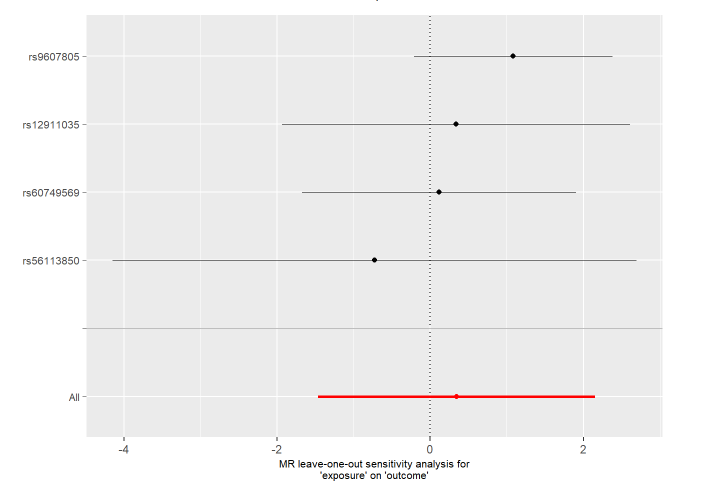

Leave one out plot demonstrating no outliers in MR analysis of Smoking cessation (n=547,219) on GD (n= 458,620).

### **Figure S5. Forest plot of variant specific inverse variance estimates for causal association between smoking behaviors and graves diseases**

Figure S5 depicts a forest plot of variant-specific inverse variance estimates, illustrating the causal association between smoking initiation, lifetime smoking, cigarettas per day, age of initiation smoking and smoking cessation on the incidence of Graves’ disease.

**Smoking Initiation**

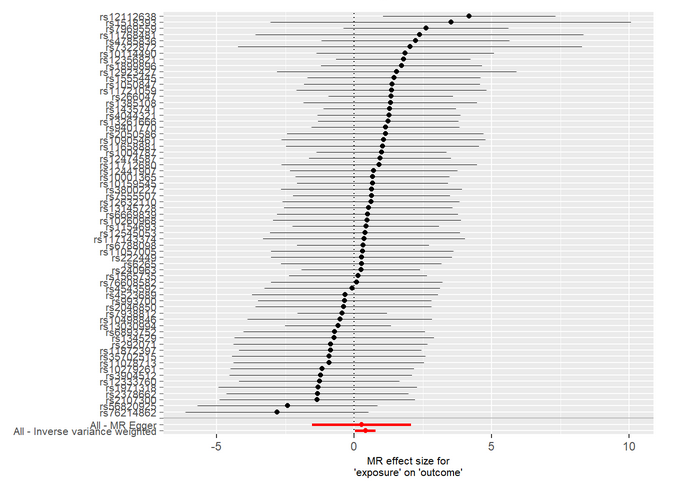

**Lifetime smoking**

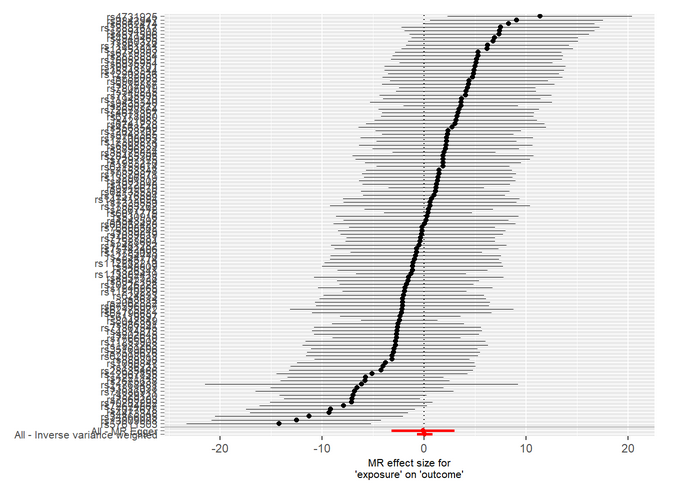

**Cigarettes per day**

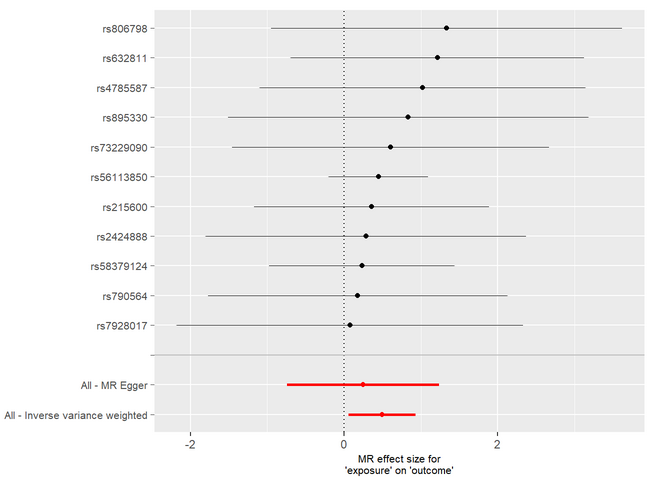

**Age of initiation smoking**

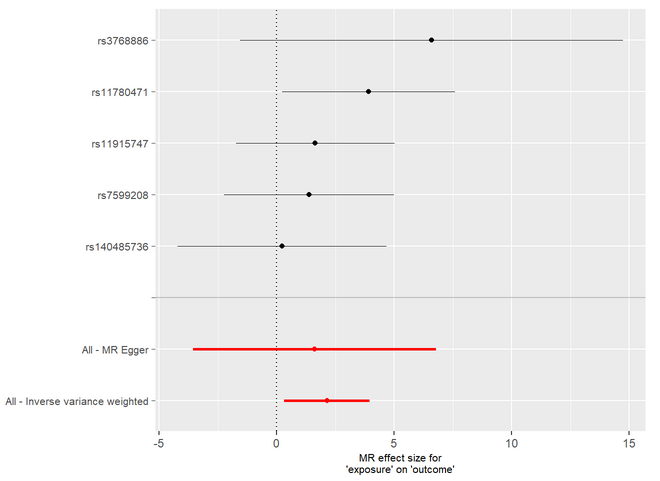

**Smoking cessation**

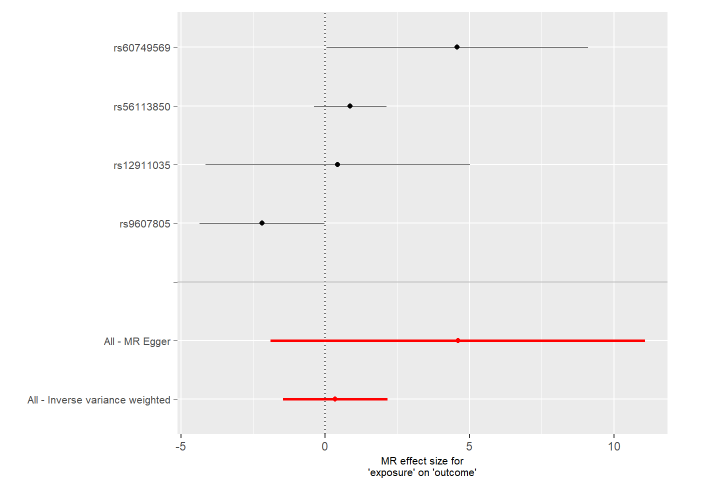

### **Figure S6. Funnel plot of causal association between Smoking behaviors and Graves’ diseases.**

Figures presents a funnel plot, illustrating the causal association between smoking initiation, lifetime smoking, cigarettas per day, age of initiation smoking and smoking cessation on the incidence of Graves’ diseases.

**Smoking Initiation**

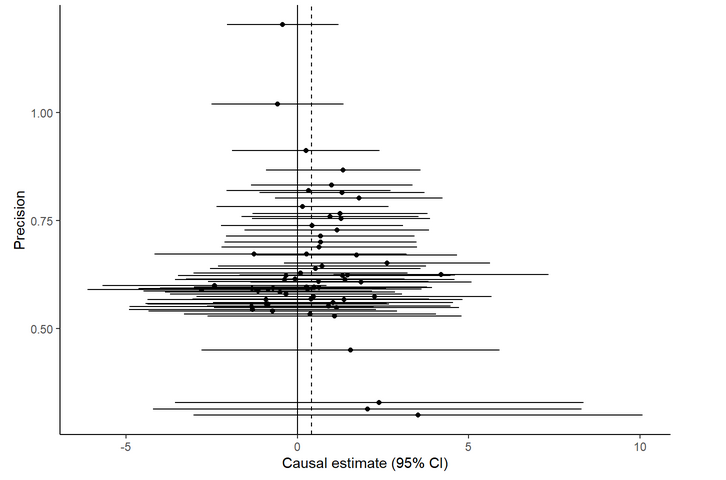

**Lifetime smoking**

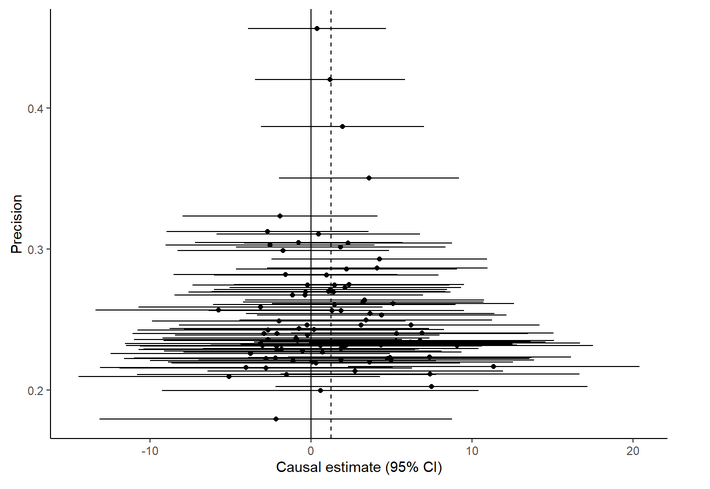

**Cigarettes per day**

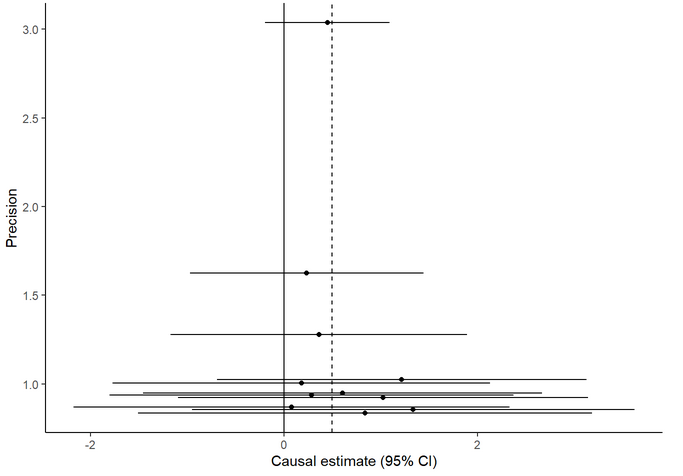

**Age of initiation smoking**

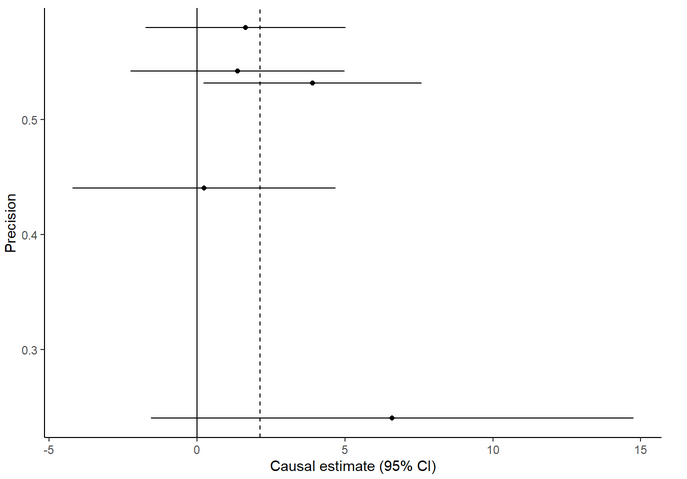

**Smoking cessation**

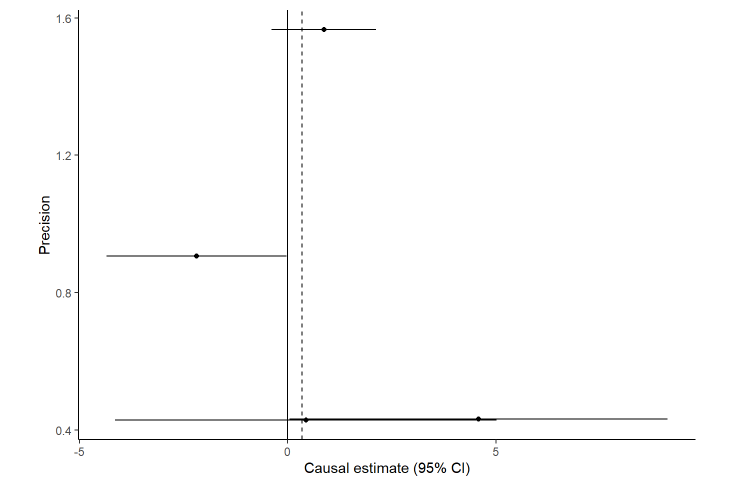

### **Figure S7. Forest plot for examine causal association between Smoking behaviors and Graves’ diseases.**

The figures below display forest plots that shows the results of different methods used to estimate the causal effect of Cigarettes per day, age of smoking initiation and smoking cessation on Graves’ disease in a Mendelian randomization analysis

**Cigarettes per day**

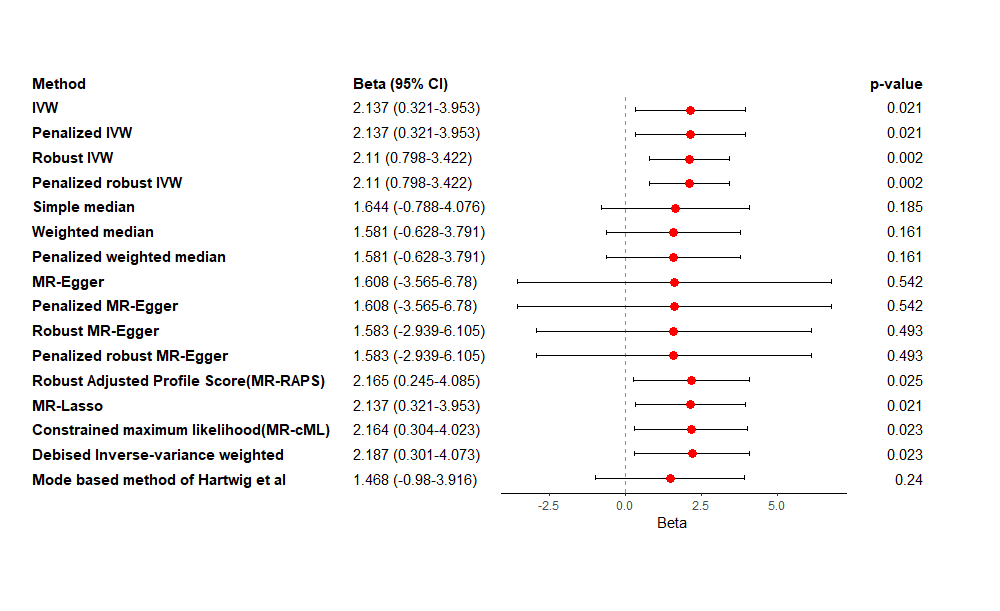

**Age of initiation smoking**

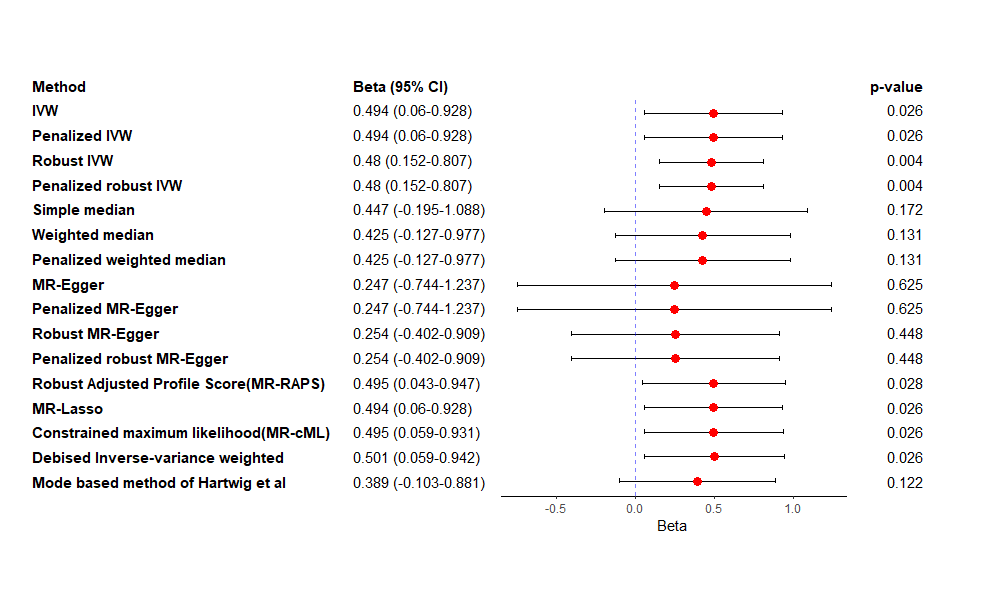

**Smoking cessation**

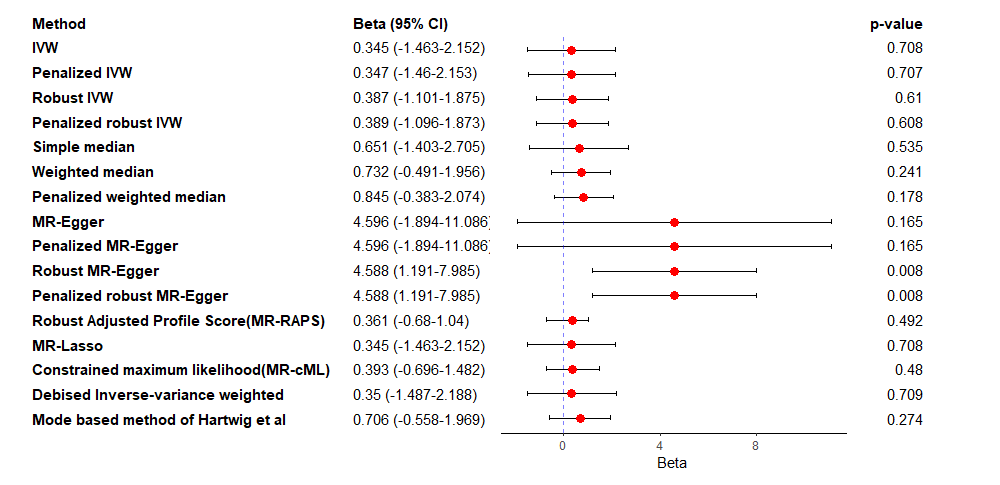

### **Figure S8. Process of instrumental variables selection**

**Smoking Initiation	Lifetime smoking	Cigarettes per day	Age of initiation smoking**

Total No. SNPs =
341,427

Total No. SNPs =
341,427

Total No. SNPs =
377,334

Total No. SNPs =
462,690

genome-wide significant**^*^** SNPs = 7,846

genome-wide significant SNPs = 447

genome-wide significant SNPs = 2,129

genome-wide significant SNPs = 10,413

After Clumping= 126

After Clumping= 93

After Clumping= 7

After Clumping= 23

After removing palindromic SNPs (harmonization) = 20

After removing palindromic SNPs (harmonization) = 7

After removing palindromic SNPs (harmonization) = 115

After removing palindromic SNPs (harmonization) = 84

After removing weak instruments = 7

After removing weak instruments = 20

After removing weak instruments = 115

After removing weak instruments = 84

After removing SNPs in HLA genes = 7

After removing SNPs in HLA genes = 20

After removing SNPs in HLA genes = 115

After removing SNPs in HLA genes = 84

After removing SNPs found in Phenoscanner= 7

After removing SNPs found in Phenoscanner= 20

After removing SNPs found in Phenoscanner= 115

After removing SNPs found in Phenoscanner= 84

After MRPRESSO = 84

After MRPRESSO = 7

After MRPRESSO = 20

After MRPRESSO = 115

After Studentized Residuals vs. Leverage Plot = 81

After Cook’s distance and residual plot = 6

After Studentized Residuals vs. Leverage Plot = 17

After Studentized Residuals vs. Leverage Plot = 112

After Q-Q Plot = 78

After Studentized residual plot = 5

After Cook’s distance and residual plot = 16

After Q-Q Plot = 109

Final number of SNPs = 5

After Cook’s distance and residual plot = 75

After DFFIT = 15

After Cook’s distance and residual plot = 107

After Studentized residual plot = 13

After DFFIT = 72

After DFFIT = 102

After single SNP plot = 65

= 89

After Studentized residual plot = 100

Final number of SNPs = 11

= 11

After single SNP plot = 11

Final number of SNPs = 65

Final number of SNPs = 95

After single SNP plot= 95

**Smoking cessation**

Total No. SNPs =
2,180

genome-wide significant SNPs = 223

After Clumping= 8

After removing SNPs in HLA genes = 8

After removing SNPs found in Phenoscanner= 8

After MRPRESSO = 8

After Cook’s distance Plot = 5

After DFFIT Plot = 4

Final number of SNPs = 4

= 11

* SNPs were selected based on a significance threshold of P < 5 × 10^−8^.
